# The trends in the uninsured rates among non-elderly adult patients hospitalized with acute pancreatitis in the US during 2004-2019

**DOI:** 10.1101/2022.05.20.22275404

**Authors:** Vivek Kumar, Ebrahim Barkoudah, David X Jin, Peter Banks, Julia McNabb-Baltar

## Abstract

**Background and Aim:** The impact of health reforms such as affordable care act (ACA) and Medicaid expansion program on health insurance coverage among acute pancreatitis (AP) patients in the United States (US) is unknown. We report the trends and forecasts for the uninsured rates among acute pancreatitis (AP) patients in the US.

**Methods:** We included non-elderly adult patients (aged 18 -64 years), hospitalized with AP in the nationwide inpatient sample database years 2004-2019. We calculated the percentage of uninsured and Medicaid patients for each year and applied joinpoint (JP) regression model to study the trends.

**Results:** The uninsured rates among patients hospitalized with AP were almost twice compared to the national average rates for all hospitalized patients. Uninsured rates were higher among Hispanic and African American races, rural location, lower income quartiles and in the southern US regions. A statistically significant decline was observed during 2013-2016 with an APC of - 12.23 (−20--4); ***p<0/01***). The decline was apparent among all racial groups, locations and income groups but not in the southern region where gap in the uninsured rates worsened compared to other geographical regions. The trends reversed more recently, and the uninsured rates surged in 2018 for the first time after 2010. In 2019, 12.5% AP patients were uninsured compared to 11.6% in 2017. The forecasts after taking unemployment rates into account showed that uninsured rates would peak in 2020 followed by a gentle decline in the following years but overall uninsured rates would remain higher compared to 2017.

**Conclusions:** The ACA and Medicaid expansion programs resulted in overall decrease in the uninsured rates, particularly among racial minorities, rural location and in the lowest income group. However, there was a surge in uninsured rate in the most recent years (2017-2019) which will continue during the next four years

---

Acute pancreatitis (AP) is one of the leading gastrointestinal causes of hospitalizations in the United States (US) with its incidence increasing^1^. In the year 2015, AP led to more than one million hospitalization days in the US, resulting in more than $2.5 billion in hospitalization charges^2^. A previous study has reported a higher incidence of uninsured patients in the year 2009-2012 compared to 2002-2005^1^. So far, no study has reported the trends in the rates of uninsured patients among AP patients. The lack of insurance has been associated with poor access to healthcare and subsequent adverse outcomes^3^. In the US, the number of uninsured patients decreased after health reforms such as the Affordable Care Act (ACA) and Medicaid expansion programs^4^. However, the impact of ACA among patients with AP is still unknown. The non-elderly (aged 18-64 years old) AP patients in the US are unique due to higher rates of alcohol and substance use, lower income groups, and a higher fraction of minority races; all these factors are also associated with a lack of health insurance in the US^5^. In this study, we aim to analyze the trends in the uninsured rates among patients hospitalized with a principal diagnosis of AP in the last 15 years, emphasizing the most recent trend changes. We examined the disparities in the health coverage among AP patients by geographical regions, race, rural/urban status, and income levels. We also project the uninsured rates among these patients for the next 4 years after adjusting for unemployment rates.

For this study, we included non-elderly adult patients, hospitalized with AP in the nationwide inpatient sample (NIS) database for the years 2004-2019^12^. We collected data on uninsured and Medicaid patients, race, rural/urban residence, income, and geographical regions for each year. We calculated the percentage of uninsured and Medicaid patients for an individual year and applied Joinpoint (JP) regression model to study the trends^6^. This program enables us to test whether an apparent change in the trend is statistically significant by using Monte Carlo Permutation method as the test of significance. The location of JP was not known prior and was estimated by the program. Annual percent change (APC) and average annual percent change (AAPC) were calculated with *‘p’* values to test if apparent changes in the trends were statistically significant. For forecasting, we applied vector autoregression (VAR) model, a multivariate time series model, and adjusted uninsured rates with the unemployment rates in the US. The unemployment rates were obtained from the US Bureau of Labor Statistics of the US department of labor ^7^. Statistical analysis was conducted using National Cancer Institute’s Joinpoint regression software and ‘R’.

The uninsured rates among patients hospitalized with AP were almost twice the national average rates for all hospitalized patients (Fig 1a). Uninsured rates were higher among Hispanic and African American races, rural locations, lower-income quartiles, and southern US regions (Table 1). The trends in the uninsured rates among AP patients mirrored the changes in the national rates, but the gap persisted (Fig 1a). Overall, the change in the uninsured rate from 2004 to 2019 was not statistically significant with an AAPC of -1 (−5.8-4.1); *p=0/7*). But a statistically significant decline was observed during 2013-2016 with an APC of -12.23 (−20--4); ***p<0/01***). A simultaneous increase in Medicaid coverage was also noticed (fig 1b). The decline was apparent among all racial groups, locations, income groups and geographical regions, however uninsured rates in the southern region remained considerably higher (Table 1). Unfortunately, trends reversed more recently, and the uninsured rates surged in 2017 for the first time after 2010. In 2019, 12.5% AP patients were uninsured compared to 11.7% in 2017. After adjusting for unemployment rates, the uninsured rates are anticipated to increase drastically in the year 2020 to 16.04% (12.8-19.3), followed by some decline in the following years 13.9(10.7-17.13) in 2021, 13.97 (10.7-17.2) in 2022 and 13.5(10.3-16.8) in 2023.

**Fig 1.**
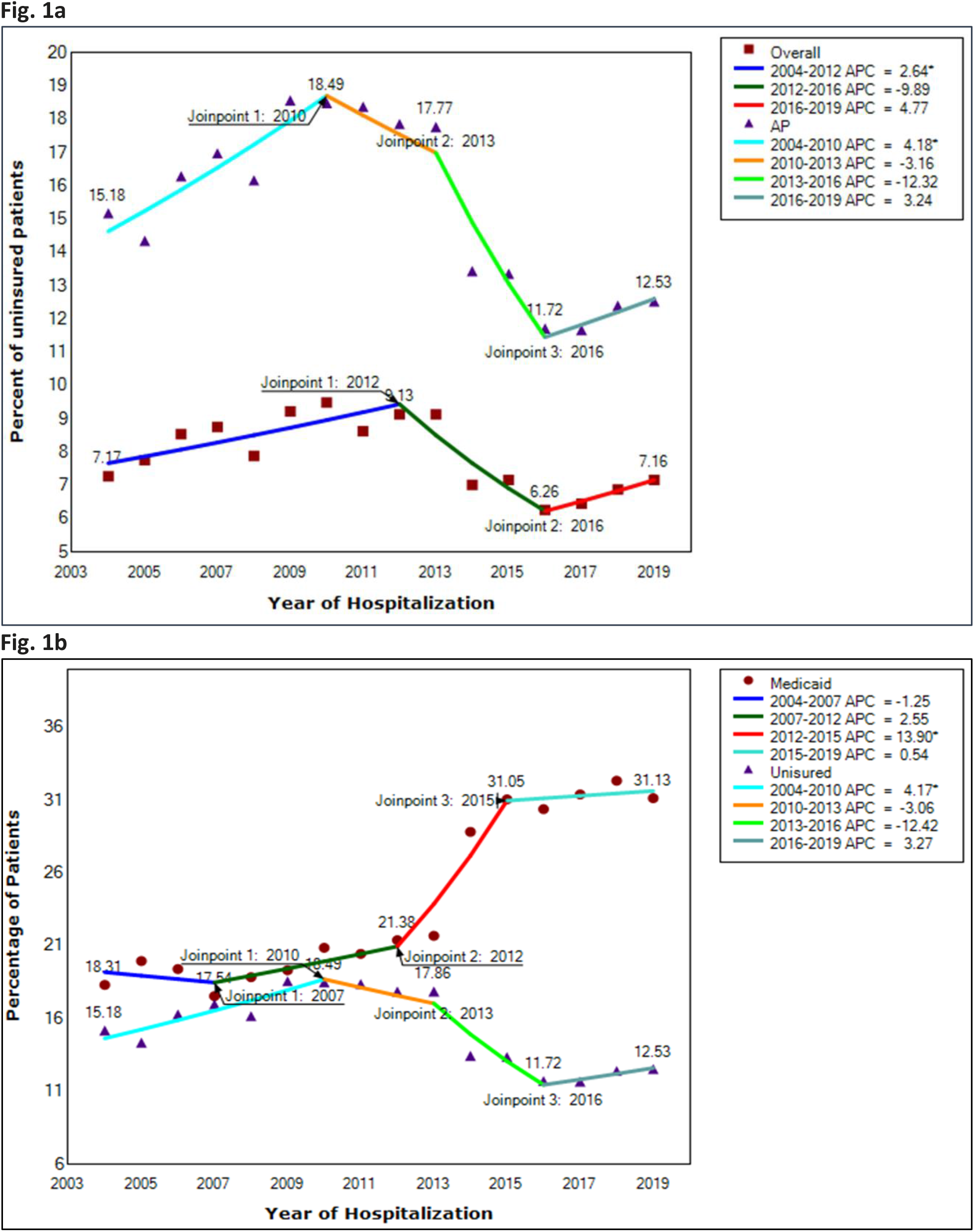
Trends in the rates of uninsured patients hospitalized with acute pancreatitis 1a. Overall, among all hospitalized patients vs AP patients. 1b. Uninsured and Medicaid insurance among patients with acute pancreatitis.

**Fig 2.**
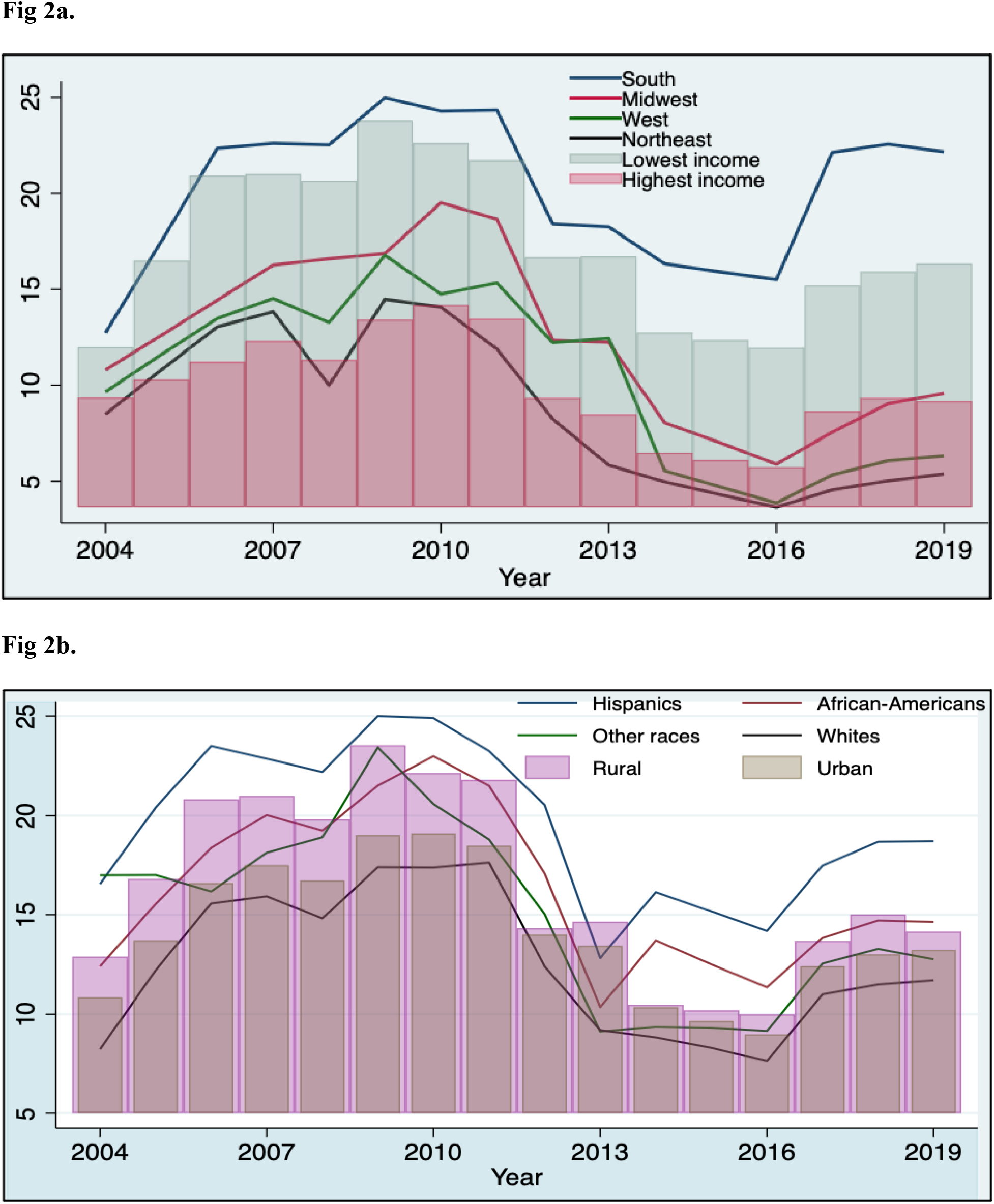
Trends in the rates of uninsured patients hospitalized with acute pancreatitis 2a. geographical regions and income groups 2b. stratified by race, and rural-urban location.

The higher uninsured rates among AP patients are due to these patients’ unique characteristics, as mentioned previously. On Joinpoint regression, we noted several periods with changes in the uninsured rates. The most prominent was a decline in the years 2013-2016. In the US, a major factor affecting the lack of insurance among the non-elderly population is unemployment^8^. While the pattern in the uninsured rates during the initial years of the study was parallel to the unemployment rates, the decline during the years 2013-2016 was out of proportion to what could be explained by declining unemployment rates alone. In fact, this corresponded to the rollout of ACA and Medicaid expansion in the US. By 2016, 33 out of 50 states had implemented Medicaid expansion. The Medicaid expansion was lower in the southern region, where only 7 out of 16 states had implemented Medicaid expansion by the year 2016^9^. This observation also explains the absence of a sharp decline among uninsured patients in the southern region, unlike other regions in the US. The higher rates of uninsured among Hispanics and African American is due to higher rates of unemployment and poverty, which is consistent with the overall rates of uninsured in the USA^10^.

Unfortunately, we noted a rising trend in the uninsured in the years 2017-2019. This trend was despite the declining rates of unemployment in the US during this time and is most likely the result of several changes made in the year 2017 to roll back ACA. These changes included waiving or delaying the features of ACA that imposed financial or regulatory burden, reduction in the funding for ACA outreach and education program. Besides, there was also discontinuation of cost-sharing reduction to insurers which led to the loss of incentives to purchase the coverage and increased the uncertainty among the eligible individuals resulting in loss of health insurance coverage ^11^.

To forecast the uninsured rates for the next three years, we adjusted for the unemployment rates. Due to the pandemic, the unemployment rates increased drastically in the US in 2020 to 6.7, almost double of 3.6 in 2019. As unemployment is a major driving factor for insurance among the non-elderly population in the US, we anticipated a drastic rise in the uninsured rates, which was found to be true. The projected rates are similar to the rates prior to implementing ACA reforms which implies that we will probably lose most of the gains in terms of health coverage that were achieved due to ACA if there are no significant changes in the policy.

The limitations of this study include the use of an administrative database using ICD-9 and ICD-10 codes which is subject to misclassification bias. Besides unemployment other factors such as patient preference may also affect the uninsured rates, which could not be analyzed due to the limitations of the database. Nevertheless, we conclude that the ACA and Medicaid expansion programs positively impacted the health insurance coverage among patients hospitalized with AP, specifically on racial minorities and low-income groups. The impact was less apparent in the southern US, where many states did not implement these reforms. Despite the strong US economy and declining unemployment rates during 2017-2019, there was a surge in uninsured patients, likely due to the changes made in the Medicaid policies. This surge is projected to continue for the next few years. Therefore, new policies and reforms are warranted for the communities at the highest risk of losing coverage-ethnic minorities and lower-income groups.

## Data Availability

All data produced in the present study are available at www.hcup-us.ahrq.gov/tech_assist/centdist.jsp.

